# Health activism, vaccine, and mpox discourse: BERTopic based mixed-method analyses of tweets from sexual minority men and gender diverse (SMMGD) individuals in the U.S.

**DOI:** 10.1101/2024.03.19.24304519

**Authors:** Yunwen Wang, Karen O’Connor, Ivan Flores, Carl T. Berdahl, Ryan J. Urbanowicz, Robin Stevens, José A. Bauermeister, Graciela Gonzalez-Hernandez

**Affiliations:** Department of Computational Biomedicine, Cedars-Sinai Medical Center, West Hollywood, CA, USA; Department of Biostatistics, Epidemiology and Informatics, Perelman School of Medicine, University of Pennsylvania, Philadelphia, PA, USA; Departments of Medicine and Emergency Medicine, Cedars-Sinai Medical Center, West Hollywood, CA, USA; Annenberg School for Communication and Journalism, University of Southern California, Los Angeles, CA, USA; Department of Family and Community Health, School of Nursing, University of Pennsylvania, Philadelphia, PA, USA

**Keywords:** mpox (monkeypox), social media, SMMGD, natural language processing, emerging infectious disease, health activism, stigma prevention, health equity

## Abstract

**Objectives:** To synthesize discussions among sexual minority men and gender diverse (SMMGD) individuals on mpox, given limited representation of SMMGD voices in existing mpox literature.

**Methods:** BERTopic (a topic modeling technique) was employed with human validations to analyze mpox-related tweets (*n* = 8,688; October 2020—September 2022) from 2,326 self-identified SMMGD individuals in the U.S.; followed by content analysis and geographic analysis.

**Results:** BERTopic identified 11 topics: health activism (29.81%); mpox vaccination (25.81%) and adverse events (0.98%); sarcasm, jokes, emotional expressions (14.04%); COVID-19 and mpox (7.32%); government/public health response (6.12%); mpox symptoms (2.74%); case reports (2.21%); puns on the virus’ naming (i.e., monkeypox; 0.86%); media publicity (0.68%); mpox in children (0.67%). Mpox health activism negatively correlated with LGB social climate index at U.S. state level, *ρ* = -0.322, *p* = 0.031.

**Conclusions:** SMMGD discussions on mpox encompassed utilitarian (e.g., vaccine access, case reports, mpox symptoms) and emotionally-charged themes—advocating against homophobia, misinformation, and stigma. Mpox health activism was more prevalent in states with lower LGB social acceptance.

**Public Health Implications:** Findings illuminate SMMGD engagement with mpox discourse, underscoring the need for more inclusive health communication strategies in infectious disease outbreaks to control associated stigma.

## Introduction

The ongoing mpox outbreak, which started in May 2022, is the first instance of human-to-human transmission in multiple non-endemic geographical areas.^1^ Formerly known as monkeypox and renamed in November 2022 to minimize stigma,^2^ mpox has caused the current outbreak to accumulate 31,010 cases and 55 deaths in the United States (U.S.) and 91,417 cases worldwide by October 26, 2023.^3^ Mpox is an infectious viral disease and a zoonosis transmitted both to and between humans: for an animal-to-human transmission, a human can contract the virus through contacting or consuming an infected animal from a range of mammal species, or through direct contact with the natural host’s blood or body fluids; between humans, mpox can be spread via direct skin-to-skin contact.^4^ Since the first identification of mpox virus among laboratory monkeys in 1957 and first report of mpox in humans in 1970, mpox has been largely confined to endemic areas in Africa, except for a small 2003 outbreak in the U.S. where transmissions occurred from infected animals to dozens of humans.^1^

During disease outbreaks, stigmatizing and discriminating against certain at-risk groups may challenge public health efforts. This occurred during the COVID-19 pandemic in the U.S., where hate crimes and violence towards people of East Asian descent surged due to perceptions attributing the pandemic to China.^5^ Similarly, sexual minority men and gender diverse (SMMGD) individuals have been battling with the so-called “gay disease” stigma attached to HIV/AIDS since the 1980s when the infection and death cases were initially reported in North America to be prevalent among SMMGD.^6^ The recent mpox outbreak, coupled with rising misinformation,^7^ stigma,^8,9^ and conspiracy theories,^10^ may, like HIV, experience a stigmatization process that leads to delayed care-seeking and further marginalization of SMMGD individuals.

In recent literature on health stigma and equity, researchers argues that the special focus of public health agencies and media on SMMGD during the mpox epidemic may fuel stigma and homophobia but not help clinically.^8,11^ However, current scientific literature about mpox poorly represents voices from this community. The literature either features expert opinions not based on original empirical research,^8^ or analyzes public opinion generally,^11^ generating results with little or no knowledge about the post author’s identity. Few studies have directly examined insights from SMMGD individuals who are arguably impacted the most by mpox epidemiologically and infodemiologically. Even in a study that did focus on mpox and this community, the analyses were performed on mpox tweets containing keywords about lesbian, gay, bisexual, transgender, queer/questioning, intersex and other sexual or gender identities (LGBTQI+), and the posts were from general users not necessarily self-identified as LGBTQI+.^9^ Not specific to mpox, one study^12^ found more negative patient experience sentiment among LGBTQI+ users than non-LGBTQI+ users, which demonstrates the importance of considering the post author’s identity when analyzing social media data. This study addresses this research gap by analyzing online mpox posts from a specific user cohort, and discussing implications of our findings for health communication tackling mpox and future disease outbreaks with an emphasis on fairness, equity, as well as stigma prevention and control.

Methodologically, this study investigates best practices when applying BERTopic^13^ on a dataset with a specific context. Topic modeling is a widely used text classification method for identifying topics in a collection of documents like social media posts, using approaches such as latent Dirichlet allocation (LDA).^14^ BERTopic^13^ is a more recent topic modeling technique that has gained popularity for its ease of interpretation and ability to leverage Hugging-Face transformers and class-based Term Frequency Inverse Document Frequency (c-TF-IDF) to create dense clusters. According to a study comparing the efficacy of four popular topic modeling approaches, namely LDA, non-negative matrix factorization (NMF),^15^ Top2Vec,^16^ and BERTopic on a Twitter dataset, BERTopic was found to perform exceptionally well and able to, like NMF, provide a more clear distinction between identified topics than LDA and Top2Vec; compared to NMF, BERTopic provides more novel insights using its embedding approach.^17^ We also investigate how to best validate and construe multi-class machine classification results through follow-up analyses.

## Methods

### Data Collection and Descriptions

The sample of this study consists of mpox-related tweets in English (*n* = 8,688) posted between October 10, 2020, and September 20, 2022, by 2,326 users who self-identified on Twitter/X as sexual minority men and gender diverse (SMMGD) individuals. The users belong to a cohort that a previous study^18^ characterized as Twitter/X users who self-identify as gay, bisexual, or men who have sex with men based on tweets and profile descriptions, with a reported accuracy rate of 85%.

For this study, we used the official Twitter Application Programming Interface (API) to collect the tweet timelines of the users mentioned above. We filtered the timeline data using mpox related keywords (i.e., “monkeypox”, “hmpxv”, “monkey pox”, and “mpox”), yielding a subset of tweets from 2,687 users who discussed mpox and were likely SMMGD. We also verified each user’s gender and sexual profile through human annotation and only retained the users with validated self-reports as SMMGD. The user validation process is described below.

### Validation of Gender/Sexual Identity Self-reports

The validation was at the user level. Firstly, we curated an evidence dataset not specific to mpox for the 2,687 users, including their profile descriptions and/or the tweets containing SMMGD gender/sexual identity keywords (see Appendix A). Secondly, informed by the evidence dataset we developed annotation guidelines (available at URL-anonymized) which were then discussed, refined, and agreed among the research team. Thirdly, two annotators double-coded 20% of users with their profile descriptions and keywords-matching tweets, and reached a substantial inter-coder reliability^19^ (kappa statistic: 0·763). Lastly, the two annotators validated the remaining 80% of the users independently. The research team including the annotators had members from the LGBTQI+ community who are familiar with LGBTQI+ terms and language use.

Through validation we were left with 2,326 SMMGD individuals and their 8,688 tweets about mpox. This yielded an 86.56% user inclusion rate.

### Data Preprocessing

To prepare the data for BERTopic modeling, we cleaned and normalized the text in the following steps in order: (1) expanding contractions (e.g., from “I’m” to “I am”); (2) translating emojis and emoticons to text; (3) removing HTTP/HTTPS links, the hashtag sign (i.e., “#”), user mentions (i.e., “@user_name”), special characters, and extra spaces; (4) lower casing; and (5) converting a list of context-specific words of multiple forms to one standard form respectively using a self-defined dictionary. For example, “monkeypox”, “mpx”, “hmpxv” and other forms of reference to mpox were converted to the standard form, “mpox.” This prevents high-frequency context-specific synonyms from occurring separately in topic representations, hence enhancing model performance by increasing the topic word diversity. See Appendix B for source words and normalized words.

Afterwards, we applied the *nltk* Python package to tokenize the text, remove stop words, and perform lemmatization.

### Implementation of BERTopic Modeling

The preprocessed posts were then passed to the BERTopic model following six steps in order: (1) transforming documents into numerical representations using *all-MiniLM-L6-v2*, a sentence-transformers model capable of capturing semantic similarity between documents; (2) using UMAP^20^ to reduce the dimensionality of input embeddings, preparing them for topic clustering; (3) applying HDBSCAN,^21^ a hierarchical clustering algorithm, to find the natural groupings or topic structure of the documents; (4) implementing *sklearn*’s CountVectorizer to convert the collection of documents to a matrix of token counts, namely, a bag-of-words representation; (5) adding to the BERTopic model a c-TF-IDF representation with additional BM-25 weighting to reduce frequent words in topic generation; and (6) fine-tuning the BERTopic representation by adjusting hyperparameters including *n_neighbors, n_components, min_dist, min_cluster_size*, and *min_samples*.

The code for this study may be made available to qualified researchers on reasonable request from the corresponding author.

### Qualitative Synthetization and Validation of Machine-generated Topic Results

Given the machine-generated topics, annotators judged the top 10 keywords and the most representative tweets of each topic to create semantic topic labels. This resulted in a k-topic label scheme. The research team read more tweets and refined the topic labels to make them more summative, preventing the labels from overfitting the example tweets reviewed.

To evaluate the quality of topic representation and the generalizability of the k-topic label scheme to unseen instances, a primary annotator read 10% of the sample (*n* = 864) and categorized them to topics without seeing the machine-assigned labels, and a secondary annotator coded for 15% of this subset to test inter-rater reliability. The topic assignments of the human annotators were individually compared to those of the machine to evaluate the BERTopic model performance.

### Testing Geographic Association with Topic Sizes

In addition, the size of the topics was compared across states (geolocations were extracted from location information in tweet metadata and user profiles using the Carmen 2.0 tool).^22^ The size of the topics indicates their relative salience, and the measure was standardized based on the total number of posts in a state before being used in geographic analyses.

To test the geolocation-based associations between online discussion patterns about mpox and the local social variables at the state level, we adopted the “LGB social climate index” aggregate by the Williams Institute at the University of California, Los Angeles^23^ to indicate the level of social acceptance of lesbian, gay, and bisexual people (LGB) of each U.S. state. We ask the research question if “health activism,” the most prominent topic in this corpus, is more prevalent in states with a higher or lower LGB social climate index. A positive correlation would suggest that health activism is more active when the social climate is more supportive; whereas, a negative correlation would suggest otherwise, namely there is more health activism in the face of oppression.

## Results

### Topic Representations

The BERTopic procedure identified 11 topics in this dataset (*n* = 8,688) and assigned one topic representation per tweet. The percent agreement rates between the machine and the primary human annotator, between the machine and the secondary human annotator, and between the two human annotators are respectively 60.1%, 60%, and 70.77%. We leveraged both machine-generated results and human interpretation to generate the topic labels.

Figure 1 shows the semantic similarity between topics. Table 1 provides the size, label, top 10 keywords, and example posts of each topic. Ranked descendingly by the topic size, the topics and their definitions were as follows.

- health activism (29.81%)
- mpox vaccination (25.81%)
- sarcasm, jokes, and emotional expressions (14.04%)
- COVID-19 and mpox (7.32%)
- government or public health response (6.12%)
- symptoms of mpox (2.74%)
- case reports (2.21%)
- mpox vaccine adverse events (0.98%)
- puns on the naming of the virus (0.86%)
- media publicity (0.68%)
- mpox in children (0.67%)

**Table 1.**
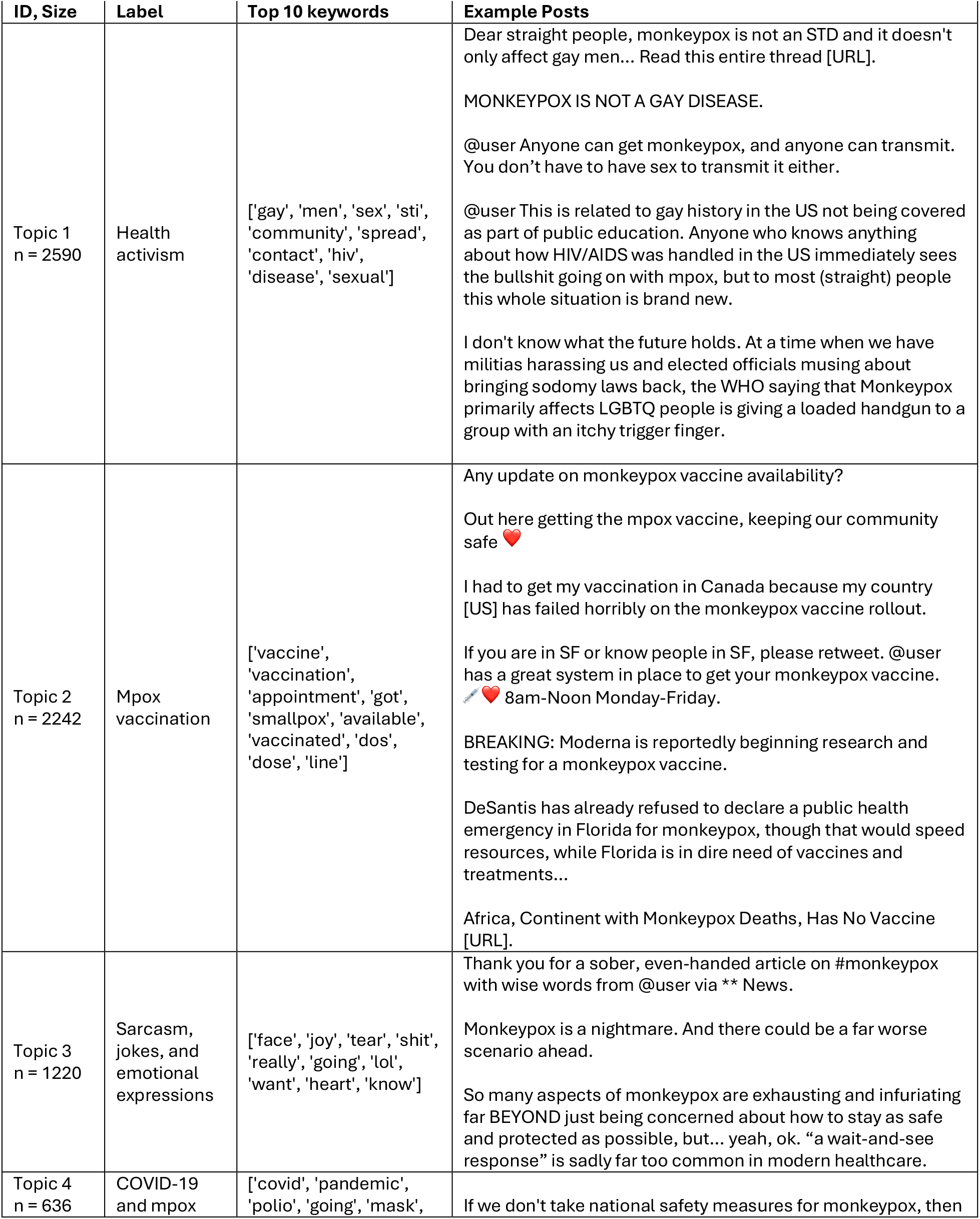
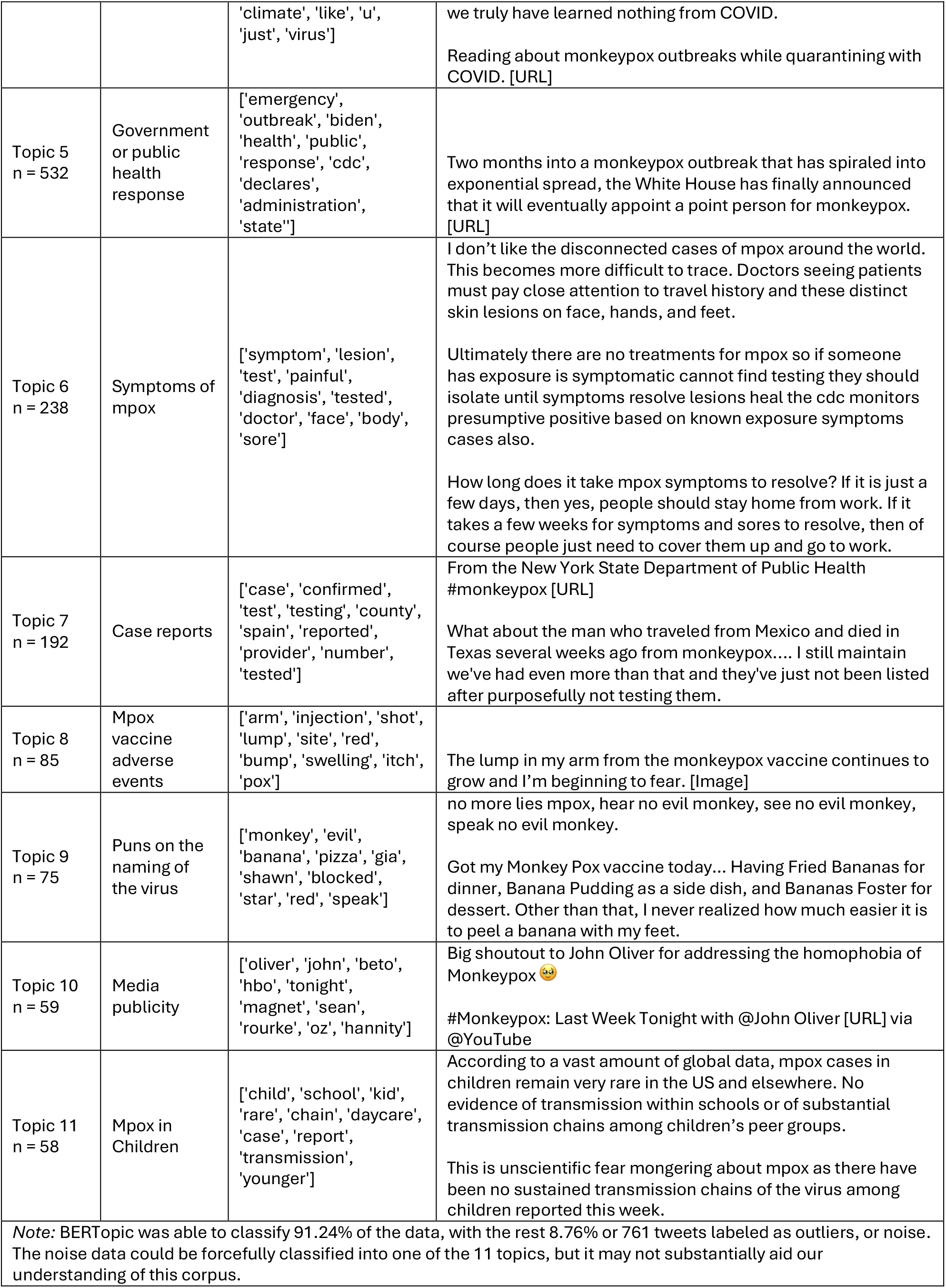
The 11-topic label scheme with the topic sizes, labels, and top keywords.

**Figure 1.**
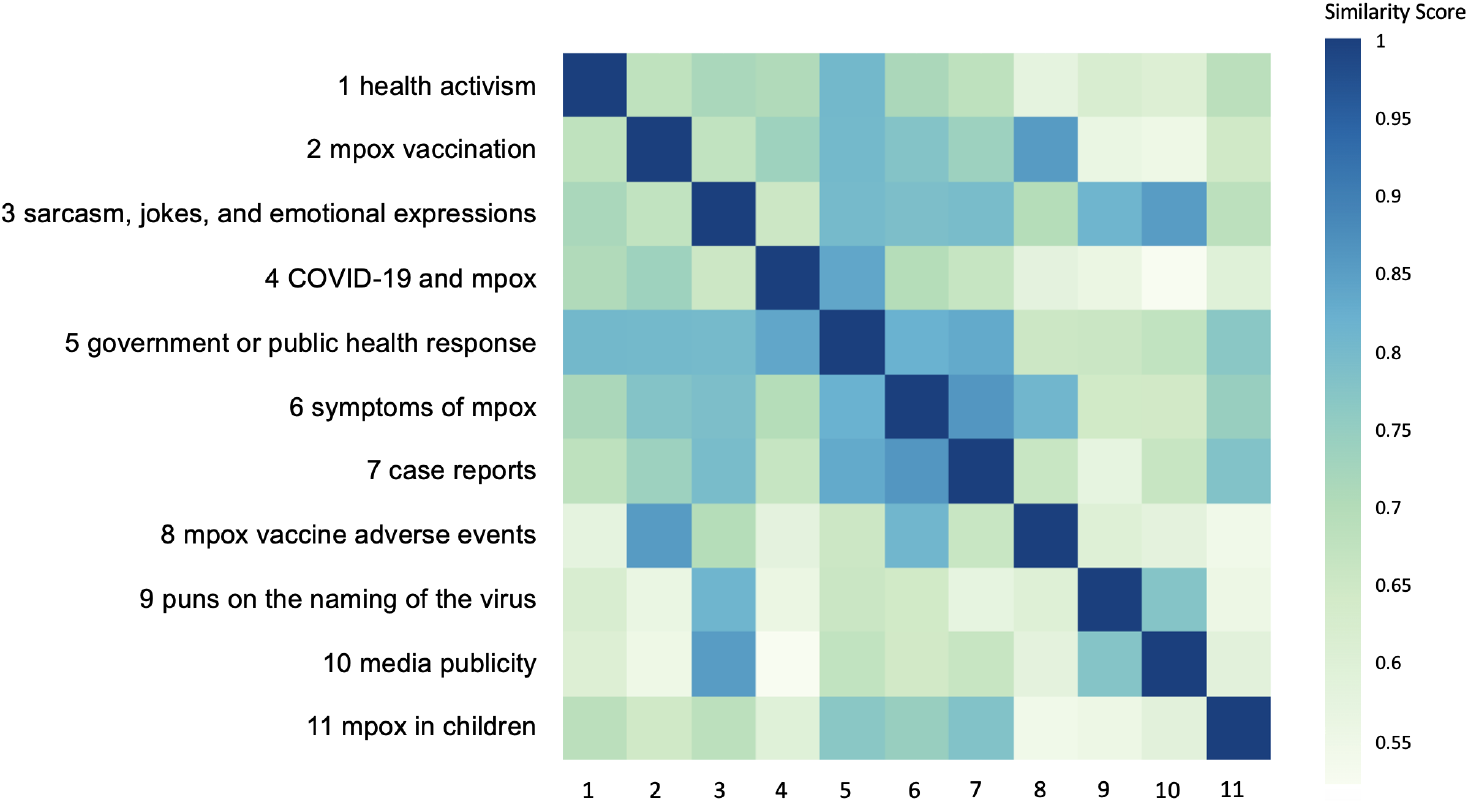
Topic similarity matrix

#### Health activism (29.81%)

Health activism refers to efforts promoting equity, fairness, and justice on a health agenda; it is aimed to counter challenges in the existing power dynamics perceived to negatively impact health communication or health outcomes.^24^ In this corpus, majority of the tweets center around health activism addressing awareness, homophobia, and health stigma of mpox.

Some posts on this topic educate the public about the mpox transmission mechanism, stressing that mpox is not a sexually transmitted infection/disease (STI/STD) and not a “gay disease.” Specifically, the epidemiological focus on the gay community is perceived by the users to have amplified an association between the gay community and mpox: “@user_name The epidemiological criteria has created impediments to testing people outside the gay community, which has ensured that the gay community remains the most visible in both criteria and counts, despite the fact that monkeypox is transmissible through a variety of kinds of contact.” Like this example, these posts sometimes engage with other users through the @mention function of Twitter/X to spark conversations.

This group of posts also reference the HIV endemic in discussing mpox. Some note the similar discursive pattern between mpox and HIV as they both tend to stigmatize the queer community. Some discuss the policy implications of existing problematic speech surrounding mpox, warning against discriminative public policies targeting SMMGD individuals.

#### Mpox vaccination (25.81%)

The second largest group of posts discuss various issues about the mpox vaccine. These posts include the importance of vaccination against mpox, inquiries or exchange of information for getting the vaccine, announcements of vaccine appointments or having gotten vaccinated, and other news on mpox vaccine.

Notably, geographical disparity regarding health resources is a recurring sub-theme in this topic, with specific geo locations such as New York City mentioned in discussing national or international vaccine shortage.

#### Sarcasm, jokes, and emotional expressions (14.04%)

The third largest group of posts focused on rhetoric and emotional expressions related to mpox, including thankfulness, fear, sadness and/or anger, and sarcasm and/or jokes.

#### COVID-19 and mpox (7.32%)

With lingering effects of COVID-19 on the public, this fourth-largest topic is characterized by discussing mpox in comparison to, or in relation to, COVID-19 in public health measures, resources etc.

#### Government or public health response (6.12%)

This group of posts start with the state or federal government declaring mpox as a public health emergency. As the outbreak develops, more tweets have been posted discussing various public health responses to the mpox outbreak, where we observe critiques on the administration.

#### Symptoms of mpox (2.74%)

This topic contains posts delineating specific symptoms (e.g., skin lesions on face, hands, and feet)—sometimes accompanied by images, and posts mentioning mpox disease symptoms as an aspect of this issue (e.g., how long it takes symptoms to resolve, the implications of symptom features for work and community transmission).

#### Case reports (2.21%)

In this topic, most tweets are about case numbers at a national, state, or city level, and there are individual cases reporting as well.

#### Mpox vaccine adverse events (0.98%)

About one percent of the data are distinguished by the machine from the second largest topic, *mpox vaccination*, to specifically discuss the adverse events related to mpox vaccine. The adverse events include lump, bump, swelling, and itchiness in the arm where the injection was received.

#### Puns on the naming of the virus (0.86%)

Despite being renamed to mpox, the term monkeypox is still referenced frequently by layman users online. This group of posts entail puns on the previous naming of the virus around “monkey.”

#### Media publicity (0.68%)

Some posts either praised or criticized or mentioned in a neutral tone such popular media as television shows for their discussions about mpox. For example, a subgroup of posts praised *Last Week Tonight with John Oliver* for addressing LGTBTQA selfcare as well as homophobia related to mpox; other posts mentioned Fox’s *The Dr. Oz Show* for its explanation on the theory of mpox’s origin.

#### Mpox in children (0.67%)

The last topic centers around if and how mpox affects children. Posts mostly stated that mpox is rare in children and no evidence suggests a transmission chain in children’s peer group. Occasionally, there were also messages about anti fearmongering or debunking of such misinformation/disinformation as queer teachers tend to spread mpox to school kids. Despite constituting the smallest group in this corpus, these posts focused on a specific vulnerable population and were worthy of close attention.

### Geographic Associations between health activism and LGB social climate

Figure 2 heatmap visualizes the state-level raw frequencies of posts. Figure 3 heatmaps geographically show the standardized topic sizes or within-state topic weights (i.e., the proportion of posts assigned to a topic out of all posts from that state, as explained in the methods section) of topic #1, “health activism” addressing awareness, homophobia, and health stigma of mpox; and topic #2, “mpox vaccination” discussions such as appointment, vaccination status, and geographical disparity in vaccine resources.

**Figure 2.**
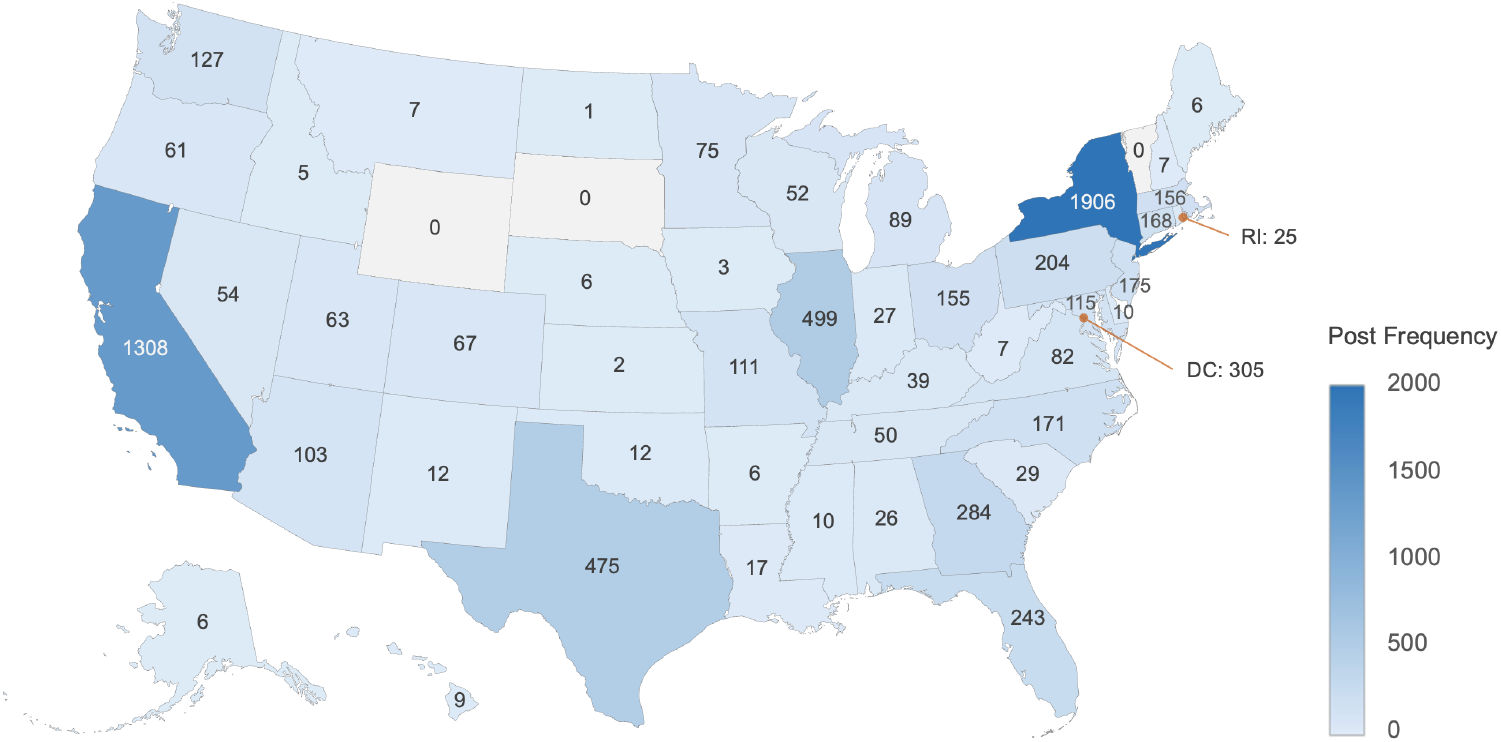
Geographic heatmap of post frequency at the U.S. Census State-level

**Figure 3.**
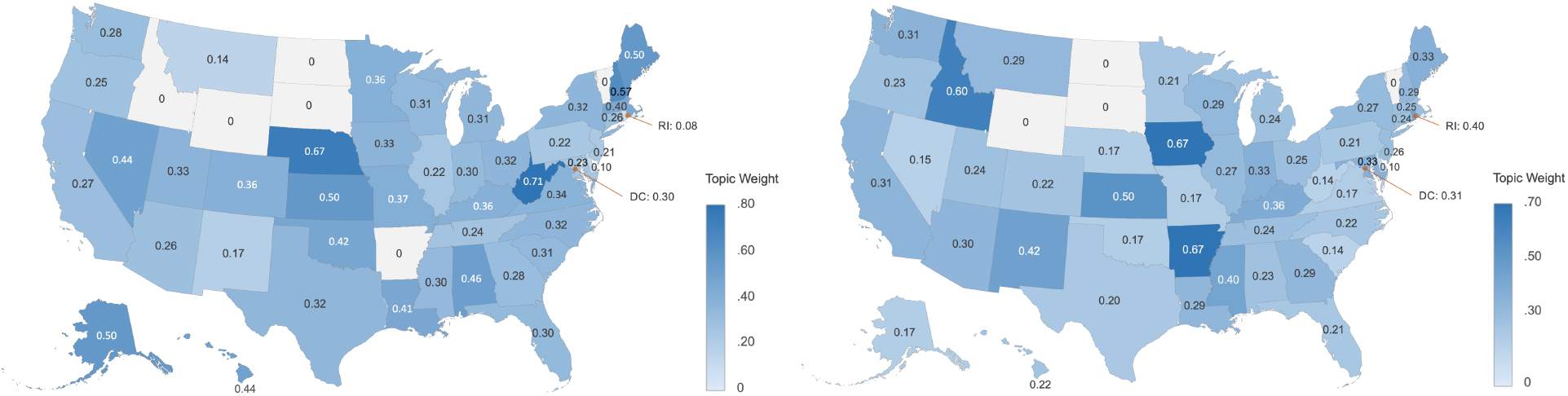
Topic #1 (health activism) and Topic #2 (mpox vaccination) weights per U.S. Census State. The topic weight is calculated as the number of topic #1 or #2 posts posted in a state divided by the total number of posts from that state.

A Spearman’s rank correlation test was performed between the state-level standardized topic size of health activism and the state-level LGB social climate index. Results indicated a negative correlation (*ρ* = -0·322, *p* = 0·031), suggesting that health activism about mpox was more active in states with less LGB social acceptance.

## Discussions and Conclusions

Public response to the mpox outbreak highlights a long-standing problem. That is, disease outbreaks often trigger health stigma, and history repeats itself by further marginalizing the communities perceived to be linked to certain diseases. Mpox is no exception.

This study combines computational and human strategies to have closely examined mpox-related online discussions among SMMGD individuals on Twitter/X. Findings fill the gap in current research, which is not inclusive of LGBTQI+ perspectives explicitly, by focusing on the discourse among SMMGD users. Based on the 11 topics we identified through BERTopic, such engaged counter-argumentation is achieved through multi-faceted mpox discussion covering health activism (29.81%), mpox vaccination (25.81%), sarcasm, jokes, and emotional expressions (14.04%), COVID-19 and mpox (7.32%), government or public health response (6.12%), symptoms of mpox (2.74%), case reports (2.21%), mpox vaccine adverse events (0.98%), puns on the naming of the virus (0.86%), media publicity (0.68%), and mpox in children (0.67%). While utilitarian content (e.g., vaccine access, case reports, mpox symptoms) constitutes a considerate part of this corpus, much of the content is emotionally charged and aimed at increasing awareness and advocating against homophobia and health stigma of mpox.

For example, compared to previous research that does not highlight SMMGD voices, our study uniquely found that SMMGD individuals opposed the narratives that “mpox is an STI/STD” and “mpox is a gay disease,” for that the popularization of such misinformation/disinformation can prevent the public from understanding the complete transmission mechanisms of mpox, challenging public health. It also risks exacerbating homophobia and health stigma by associating a disease to the community. Thus, in addition to highlighting social media users’ disclosure of attention on mpox prevention and control, our study also sheds light onto the social implications of disease outbreaks, calling for health practitioners and researchers to critically reflect on the way in which we collectively frame and narrate mpox and future infectious diseases to prevent and mitigate resulting biases and oppression.

Our findings provide implications for health communication tackling mpox and future disease outbreaks with an emphasis on fairness and equity. Corroborating previous research that found links between HIV and homophobia,^6^ and between COVID-19 and anti-Asian racism,^5^ this study found that mpox is not only linked to homophobia but also health activism. Our geographic analyses suggest that health activism was more likely to be motivated by oppression rather than accepting social climate in the U.S. at a census state level; future research can continue to test our geographic finding.

Meanwhile, users’ affect-enriched praises and criticisms of administration and media regarding their response to mpox evidence how social media enables political/media engagement for social change. From the perspectives of non-profit health organizations and government agencies, future public health efforts could use real-time monitoring of social media content to support health activism and strategically plan anti-stigma messages to reduce, control, and prevent misinformation as well as stigmatizing messages during disease outbreaks, a time of crisis that is prone to fear-mongering, targeted blame, and othering.

Methodologically, this study contributes to the current literature in three aspects. First, we show how to leverage a previously published dataset with a cohort of our interest to collect and validate another dataset for our own use case. Second, we detail how to apply BERTopic for categorizing the themes arising from a user cohort’s posts, amplifying their voices by synthesizing unstructured text data into a structured multi-topic scheme for easier interpretation. Third, we validate BERTopic results as a multi-class classification task via three-way comparisons: (i) human annotator #1 vs. human annotator #2, (ii) machine vs. human annotator #1, and (iii) machine vs. human annotator #2. This validation strategy considers the inherent difficulty of multi-class classification in machine learning, and assesses the performance of the model after adjusting for how well human annotators achieve inter-rater reliability.

Regarding study limitations, due to the large sample size, we only validated machine-generated topic results on a fraction of data, although it was sufficient to lend credibility to large-scale automatic analysis on the whole dataset. Meanwhile, some posts may belong to more than one topic while BERTopic assigns the most prominent topic to each post. In future research, we will explore the best ways of analyzing data when more than one topic is assigned to each post.

In conclusion, this study uniquely illuminates the response of a large group of sexual minority men and gender diverse individuals to the mpox outbreak and to the public reaction to mpox. BERTopic modeling validated by human annotators reveals 11 topics themed around (1) health activism against to the misinformation, disinformation, and stigma associated with mpox, (2) utilitarian content such as information exchange on mpox vaccine, disease symptoms, and public health measures, and (3) affective, rhetoric, and political expressions to the mpox outbreak and to the related public reactions. All these are key to a global public health paradigm with enhanced equity, inclusion, and fairness. Through this study, we have gathered a previously neglected segment of public opinions and performed “social listening”^25^ by analyzing social media big data using natural language processing. Results of this study can inform medical experts and health researchers about the best communicative practices around the ongoing mpox outbreak, as well as when the next disease outbreak hits mankind.

## Data Availability

All data produced in the present study are available upon reasonable request to the authors.

## Acknowledgements

Research reported in this work was partially supported by the National Library of Medicine of the National Institutes of Health under award number R01LM011176. The content is solely the responsibility of the authors and does not necessarily represent the official views of the National Institutes of Health.

### Appendix A

Regular Expressions Used by Klein et al. (2022) to Detect Sexual/Gender Identities in Twitter Profiles and Tweets

REGEXES_TWEETS = [

r’\b(dm|send|me|my|i|i\W?m)\b.*#(truvadawhore|gaytop|gayvers|teambottom|teamvers|b areback|teamtop|gaybottom)\b’,

r’\b(i\W?m|i\s+am|as)\s+(?!not)(\S+\s+)?a\s+(queer|bi\W?sexual|bi|gay)\s+(man|guy|male| dude|boy)\b’,

r’\bmy\s+fellow\s+(queer|bi\W?sexual|bi|gay)\s+(men|guys|males|dudes|boys)\b’,

r’\b(queer|bi\W?sexual|bi|gay)\s+(man|men|guy|guys|male|males|dude|dudes|boy|boys)\ W+like\s+(me|myself)\b’

]

REGEXES_PROFILE = [

r’\b(queer|bi\W?sexual|bi|gay)\b.*\b(man|guy|male|dude|boy|he|him|his|husband|father|d ad)\b’,

r’\b(man|guy|male|dude|boy|he|him|his|husband|father|dad)\b.*\b(queer|bi\W?sexual|bi|g ay)\b’,

r’#(truvadawhore|gaytop|gayvers|teambottom|teamvers|bareback|teamtop|gaybottom)\b’

]

### Appendix B

Words Normalization Rules

(Source Words: Normalized Word)

monkeypox, monkey pox, mpox, mpx, hmpxv: mpox

antivaxxer, antivaxxers: antivaxxer

vaccinated, vaxxed, vaxed: vaccinated

vaccine, vaccines, vax, vaxx, vaxs: vaccine

outbreak, out break: outbreak

sti, stis: sti

std, stds: std

covid, covid 19, covid19: covid

## References

1. The Lancet Infectious Diseases. Monkeypox: A neglected old foe. Published 2022. Accessed December 15, 2023. https://www.thelancet.com/journals/laninf/article/PIIS1473-3099(22)00377-2/fulltext

2. World Health Organization. WHO recommends new name for monkeypox disease. Published 2022. Accessed December 15, 2023. https://www.who.int/news/item/28-11-2022-who-recommends-new-name-for-monkeypox-disease

3. U.S. Centers for Disease Control and Prevention. 2022-2023 outbreak cases and data. Published 2022. Accessed December 15, 2023. https://www.cdc.gov/poxvirus/mpox/response/2022/index.html

4. Awadi, S. et al. Human Monkeypox virus in the shadow of the COVID-19 pandemic. J. Infect. Public Health. 2023;16, 1149–1157. 10.1016/j.jiph.2023.05.013

5. Hong, T. et al. Effects of #coronavirus content moderation on misinformation and anti-Asian hate on Instagram. New Media & Society. 2023;4, 14614448231187529. 10.1177/14614448231187529

6. Alonzo, A. A. & Reynolds, N. R. Stigma, HIV and AIDS: An exploration and elaboration of a stigma trajectory. Social Science & Medicine. 1995;41, 303–315. 10.1016/0277-9536(94)00384-6

7. Ennab, F. et al. Monkeypox outbreaks in 2022: Battling another “pandemic” of misinformation. International Journal of Public Health. 2022; 67; 1605149. 10.3389/ijph.2022.1605149

8. Bragazzi, N. L., Khamisy-Farah, R., Tsigalou, C., Mahroum, N., Converti, M. Attaching a stigma to the LGBTQI+ community should be avoided during the monkeypox epidemic. Journal of Medical Virology. 2022;95, e27913. 10.1002/jmv.27913

9. Movahedi Nia, Z. et al. Mpox panic, infodemic, and stigmatization of the two-spirit, lesbian, gay, bisexual, transgender, queer or questioning, intersex, asexual community: Geospatial analysis, topic modeling, and sentiment analysis of a large, multilingual social media database. Journal of Medical Internet Research. 2023;25, e45108. 10.2196/45108

10. Zenone, M. & Caulfield T. Using data from a short video social media platform to identify emergent monkeypox conspiracy theories. JAMA Network Open. 2022;5, e2236993–e2236993. 10.1001/jamanetworkopen.2022.36993

11. Keum, B. T., Hong, C., Beikzadeh, M., Cascalheira, C. J. & Holloway, I. W. Mpox stigma, online homophobia, and the mental health of gay, bisexual, and other men who have sex with men. LGBT Health. 2023;10, 408–410. 10.1089/lgbt.2022.0281

12. Hswen, Y. et al. Investigation of geographic and macrolevel variations in LGBTQ patient experiences: Longitudinal social media analysis. Journal of Medical Internet Research. 2020;22, e17087. 10.2196/17087

13. Grootendorst, M. BERTopic: Neural topic modeling with a class-based TF-IDF procedure. arXiv Preprint. 2022. https://arxiv.org/pdf/2203.05794.pdf

14. Blei, D., Ng, A. & Jordan M. Latent Dirichlet allocation. Advances in Neural Information Processing Systems. 2001;14. https://proceedings.neurips.cc/paper/2001/file/296472c9542ad4d4788d543508116cbc-Paper.pdf

15. Lee, D. & Seung, H. S. Algorithms for non-negative matrix factorization. Advances in Neural Information Processing Systems. 2000;13, 1–7. https://proceedings.neurips.cc/paper_files/paper/2000/file/f9d1152547c0bde01830b7e8bd60024c-Paper.pdf

16. Angelov, D. Top2Vec: Distributed representations of topics. arXiv Preprint. 2020;2008.09470. http://arxiv.org/pdf/2008.09470v1

17. Egger, R. & Yu, J. A topic modeling comparison between LDA, NMF, Top2Vec, and BERTopic to demystify Twitter posts. Frontiers in Sociology. 2022;7, 886498. 10.3389/fsoc.2022.886498

18. Klein, A. Z., Meanley, S., O’Connor, K., Bauermeister, J. A. & Gonzalez-Hernandez, G. Toward using Twitter for PrEP-related interventions: An automated natural language processing pipeline for identifying gay or bisexual men in the United States. JMIR Public Health and Surveillance. 2022;8, e32405. 10.2196/32405

19. Viera, A. J. & Garrett, J. M. Understanding interobserver agreement: The kappa statistic. Fam Med. 2005;37(5), 360–363.

20. McInnes, L., Healy, J. & Melville, J. UMAP: Uniform manifold approximation and projection for dimension reduction. arXiv Preprint. 2018;1802.03426. https://arxiv.org/pdf/1802.03426.pdf

21. Campello RJ, Moulavi D, Sander J. Density-based clustering based on hierarchical density estimates. In Pacific-Asia Conference on Knowledge Discovery and Data Mining. 2013;160-172. Berlin, Heidelberg: Springer Berlin Heidelberg. 10.1007/978-3-642-37456-2_14

22. Zhang J, DeLucia A, Dredze M. Changes in tweet geolocation over time: A study with Carmen 2.0. In Proceedings of the Eighth Workshop on Noisy User-generated Text (W-NUT 2022). 2022;1-14. https://aclanthology.org/2022.wnut-1.1

23. Hasenbush A, Flores A, Kastanis A, Sears B, Gates G. The LGBT divide: A data portrait of LGBT people in the midwestern, mountain & southern states. UCLA Williams Institute. Published in 2014. https://escholarship.org/uc/item/17m036q5

24. Zoller HM. Health activism: Communication theory and action for social change. Communication Theory. 2005;15, 341–364. 10.1111/j.1468-2885.2005.tb00339.x

25. Cole-Lewis H, Pugatch J, Sanders A, Varghese A, Posada S, Yun C, Schwarz M, Augustson E. Social listening: a content analysis of e-cigarette discussions on Twitter. Journal of Medical Internet Research. 2015;17, e243. 10.2196%2Fjmir.4969

